# Automated analysis of digital medical images in cervical cancer screening: A systematic review

**DOI:** 10.1101/2024.09.27.24314466

**Authors:** Leshego Ledwaba, Rakiya Saidu, Bessie Malila, Louise Kuhn, Tinashe E.M. Mutsvangwa

## Abstract

**Background:** Cervical cancer screening programs are poorly implemented in LMICs due to a shortage of specialists and expensive diagnostic infrastructure. To address the barriers of implementation researchers have been developing low-cost portable devices and automating image analysis for decision support.

However, as the knowledge base is growing rapidly, progress on the implementation status of novel imaging devices and algorithms in cervical cancer screening has become unclear. The aim of this project was to provide a systematic review summarizing the full range of automated technology systems used in cervical cancer screening.

**Method:** A search on academic databases was conducted and the search results were screened by two independent reviewers. Study selection was based on eligibility in meeting the terms of inclusion and exclusion criteria which were outlined using a Population, Intervention, Comparator and Outcome framework.

**Results:** 17 studies reported algorithms developed with source images from mobile device, viz. Pocket Colposcope, MobileODT EVA Colpo, Smartphone Camera, Smartphone-based Endoscope System, Smartscope, mHRME, and PiHRME. While 56 studies reported algorithms with source images from conventional/commercial acquisition devices. Most interventions were in the feasibility stage of development, undergoing initial clinical validations.

**Conclusion:** Researchers have proven superior prediction performance of computer aided diagnostics (CAD) in colposcopy (>80% accuracies) versus manual analysis (<70.0% accuracies). Furthermore, this review summarized evidence of the algorithms which are being created utilizing portable devices, to circumvent constraints prohibiting wider implementation in LMICs (such as expensive diagnostic infrastructure). However clinical validation of novel devices with CAD is not yet implemented adequately in LMICs.

## Introduction

Cervical cancer remains a critical public health issue, particularly in low– and medium-income countries (LMICs), where it is the second highest cause of mortality among women [1]. The World Health Organization (WHO) highlighted that of the estimated 604,000 new cases in 2020, 90% were in LMICs, with South Africa experiencing disproportionately higher rates partly due to the high prevalence of HIV [2,3]. This is underscored by the age-standardized mortality rates in South Africa being 7 to 10 times higher than in developed countries [5]. A notable disparity exists between LMICs and developed countries in cervical cancer incidence and mortality rates, largely attributed to the lack of organized population screening in resource-limited settings [1,5].

The preventability of cervical cancer through early detection of pre-malignant lesions underscores the importance of effective screening programs. However, in LMICs, common challenges such as limited specialist availability and the high cost of diagnostic equipment, particularly in colposcopy and cytology, impede the implementation of effective screening strategies [9]. This has led to a high burden of preventable cervical cancer cases [5].

Addressing these challenges, researchers are developing low-cost, portable equipment, some incorporating automated image analysis for diagnostic support [6]. These technological advancements hold promise in mitigating the screening challenges by offering cost-effective and accessible solutions. However, the landscape of such automated technology systems for cervical cancer screening is not well-understood, with existing literature being fragmented and lacking in comprehensiveness [6]. Review literature often misses emerging technologies, especially those incorporating automated digital image analysis algorithms across various optical screening domains.

### Contextualizing Cervical Cancer Screening Challenges in LMICs

The cervical cancer screening landscape in LMICs is marked by significant challenges that impede the effective prevention and management of this disease. Maimela et al. [3] reported that in South Africa, the time from cytology (pap smear) to colposcopy can exceed six months, leading to delayed care and increased mortality from preventable cervical cancer. This critical delay in treatment is attributed to the limited availability of colposcopy services at the primary healthcare level, constrained by limited budgets for specialized clinical equipment and a scarcity of specialist gynaecologists [1, 3]. As a result, colposcopy service provision is often confined to tertiary-level facilities, creating significant access barriers.

The constraints in healthcare service delivery, spanning cytology, colposcopy, and histopathology, are multifaceted. In cytology and histopathology, the reliance on manual slide analysis by cytotechnologists or pathologists is particularly challenging. This process is time-consuming and prone to reader variability errors [10, 11, 12], which compounds the difficulties faced in implementing effective cervical cancer screening programs in LMICs as recommended by the WHO [13].

Commercially available cytology scanners like ThinPrep® Imaging System and BC Focal Point™ GS Imaging System, and histopathology scanners like Aperio are known for their effectiveness. However, their high purchase and operational costs [15, 16, 17] make these effective technologies impractical for widespread use in mass screening programs in LMICs. This financial barrier is a significant challenge not only in the realm of cytology and histopathology but extends across all cervical cancer screening domains.

In response to these challenges, researchers have developed inexpensive portable equipment in colposcopy [18], cytology [16], and histopathology [17], some of which have incorporated automated image analysis to provide diagnostic decision support. The developers of MobileODT [23], Pocket colposcope [24], and Gynocular [25] have recently announced on their respective websites that research into building automation functionality into the companies’ product portfolio has begun. These innovations address not only the challenges of specialist availability but also the issue of equipment costs.

Systematic reviews, such as those by Rossman et al. [19], Allanson et al. [20], and others have provided valuable insights into digital interventions and the accuracy of specialists’ interpretations using various imaging devices. However, many of these reviews have overlooked the broader application and potential of comprehensive decision support technology systems, which include a combination of acquisition devices and computer aided diagnosis (CAD) algorithms [21–22]. This omission signifies a gap in fully understanding the landscape of emerging technologies, particularly those still in development or undergoing proof-of-concept investigations. Moreover, systematic reviews of CAD algorithms in colposcopy, cytology, and histopathology, such as those by Fernandes et al. [26], Conceicao et al. [27], and de Matos et al. [28], have often focused on model development in a single domain in isolation. This narrow focus leaves out the contextualization of acquisition devices suitable for large-scale screening programs in LMICs that could be paired with CAD algorithms.

Our systematic review aims to address this research gap by covering three domains (colposcopy, cytology, and histopathology) in which CAD algorithms are applied. This multi-domain approach allows for the identification of all automated technologies relevant to optical cervical cancer screening, thus making it more comprehensive than similar prior works. Unlike earlier reviews that only covered model development, this review includes a holistic view of the technology system in order to address both infrastructure and specialist availability challenges in LMICs. This could aid clinical researchers and policymakers in making informed decisions about integrating these technologies into community screening programs or clinical trials, particularly in LMIC contexts.

## 1. Methodology

The Preferred Reporting Items for Systematic Reviews and Meta-Analyses (PRISMA) guidelines were followed in executing the methodology [29]. Hence, a Prospero record (ID: CRD42022303174) was registered defining a clear search strategy based on PICO, which was applied on five main academic databases (PubMed, Scopus, EBSCOhost, Web of Science and Google Scholar). The academic database search was conducted between January – May 2022. This combination of databases is inclusive of American, European, and African publications; thus, ensuring that relevant research from LMICs was not overlooked throughout the search. Furthermore, Google search and Google Scholar (first 10 pages) were used to search grey literature and additional articles discovered from reference lists. Grey literature search was conducted between January – June 2022. The default Google search engine language setting was English and geography setting was South Africa. Searches were also conducted on the following patent search engines: Espacenet, Patent Public Search, Japan Patent Office, Yandex, and Baidu. Patent searches were conducted between July – August 2023.

### PICO framework

The search philosophy was developed using key words and phrases formulated according to the Population (P), Intervention (I), Comparator (C), Outcome (O) framework [30]. The detailed PubMed search strategy with the number of search hits returned is presented in Supplementary Table 1.

### Inclusion and exclusion criteria

Inclusion and exclusion criteria were defined during registration of the review for each component of the PICO framework, then used in study selection during screening.

For the Population/Participants component, studies involving both expert clinicians, such as gynaecologists, colposcopists, cytologists, histopathologists, as well as generalist healthcare professionals such as nurses at primary healthcare centres were included. The study subject had to be women receiving cervical cancer screening services. Studies with women who had been confirmed to have invasive cancer were excluded because those studies focused on staging interventions which were outside the scope of this systematic review.

For the Intervention component, studies investigating CAD-enabled cervical cancer screening technology systems were included. The relevant optical acquisition devices were those used to perform colposcopy, micro-endoscopy, Visual Inspection using Acetic Acid (VIA), or Visual Inspection using Lugol’s iodine (VILI); cytology; and histopathology. Thus, studies applying radiographical or ultrasonic medical imaging modalities, such as PET, CT, MRI, X-ray, and ultrasound were excluded because the images from these modalities are not used for cervical cancer screening. Studies using non-image datasets were excluded because the focus of this research was on image analysis.

For the Comparator component, studies that were included were those that measured the performance of automated interventions against the following comparators: 1) histopathological results 2) HPV results, 3) cytology expert judgement, 4) colposcopy expert judgement and/or 5) alternative state-of-the-art CAD algorithms. Studies without ground truth or alternative comparators were excluded since such studies preclude the determination of the effectiveness of proposed intervention(s).

For the Outcome component, studies whose algorithms achieved segmentation of objects/regions and/or the algorithms discriminating between binary classes or multiple classes were included. Studies conceptualizing CAD algorithms without implementation of said CAD algorithms on an image-based dataset were excluded, because those studies would not have had proven outcomes relevant to this systematic review. Thus, review papers and other systematic reviews were excluded from analysis. However, systematic review publications played a crucial role in shaping the research justification and identifying any critical interventions that the initial search might have overlooked, thus reinforcing this review’s foundation and comprehensiveness.

This systematic review excluded studies whose full text was not accessible due to university subscription constraints or lack of responses from authors who were contacted requesting access to full text papers.

### Screening and Data extraction

Two reviewers worked independently to screen 566 publications, critically appraising the titles and abstracts of studies, using the above inclusion and exclusion criteria, then appraising full text articles. A third reviewer made the final decision when there were conflicting decisions. Based on inclusion criteria, 73 articles were deemed eligible for data extraction. These included articles are listed in Supplementary Table 2. Figure 1 summarizes the selection process using the PRISMA flow diagram.

**Figure 1:**
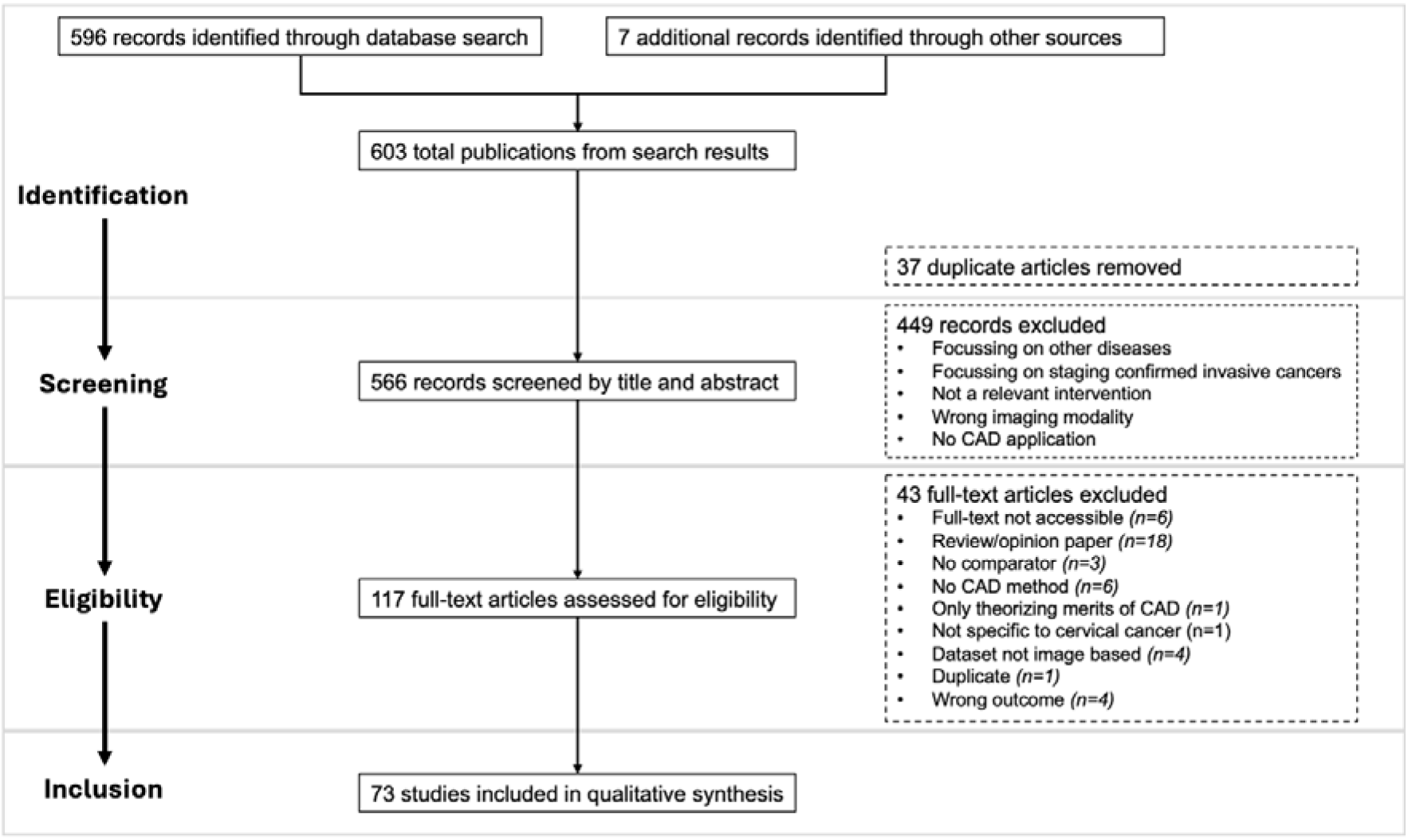
Flow of studies through different phases of systematic review.

Data extraction and study quality assessment tools (adapted from Cincinnati Children’s LEGEND guideline and JBI critical appraisal checklist for diagnostic test accuracy studies) were developed to minimise risk of bias.

## 2. Results

### Population analysis

The study methodologies of 49 authors contained information about specialists and women participating in the research. In contrast, 24 authors did not divulge information about the women participating in the research, but only mentioned the type of clinicians involved. N=30 studies included women from LMICs as the target population in the methodology (Figure 2 shows the geographical breakdown of populations). Specialists performed the screening examination for the majority of the studies (n= 47). Two studies, Cholkeri-Singh et al. [31] and Peterson et al. [32] reported nurse participation in the application of their automated colposcopy interventions.

**Figure 2:**
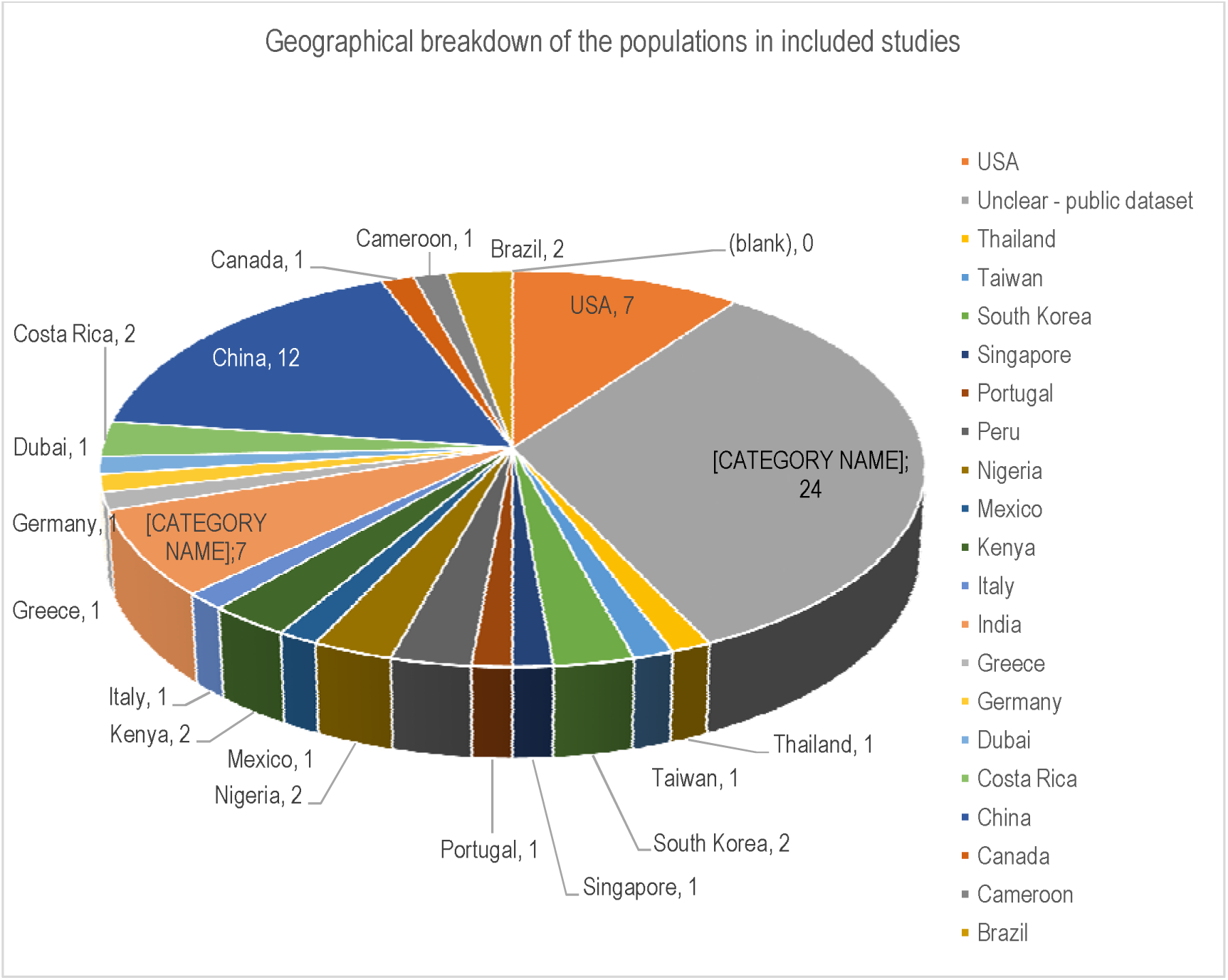
Geographical breakdown of populations in the included studies.

### Intervention analysis

The intervention analysis involved the development of a tabulated data narrative tool, which was populated with information about the interventions reported in *included*studies. The narrative tool was inspired by a review paper by Fernandes et al. [26] which presented numerous study datapoints by using a pipeline framework of the main CAD tasks involved in the development of algorithms for colposcopy domain. Based on the research question in this systematic review, there was motivation to understand how a pipeline for cytology that was outlined by Conceicao et al. [27], and a pipeline for histopathology that was outlined by de Matos et al. [28] differed from the colposcopy pipeline. This analysis approach resulted in extensive information that needed sorting and structuring; thus, the need for a narrative tool presented in Table 1.

**Table 1:**
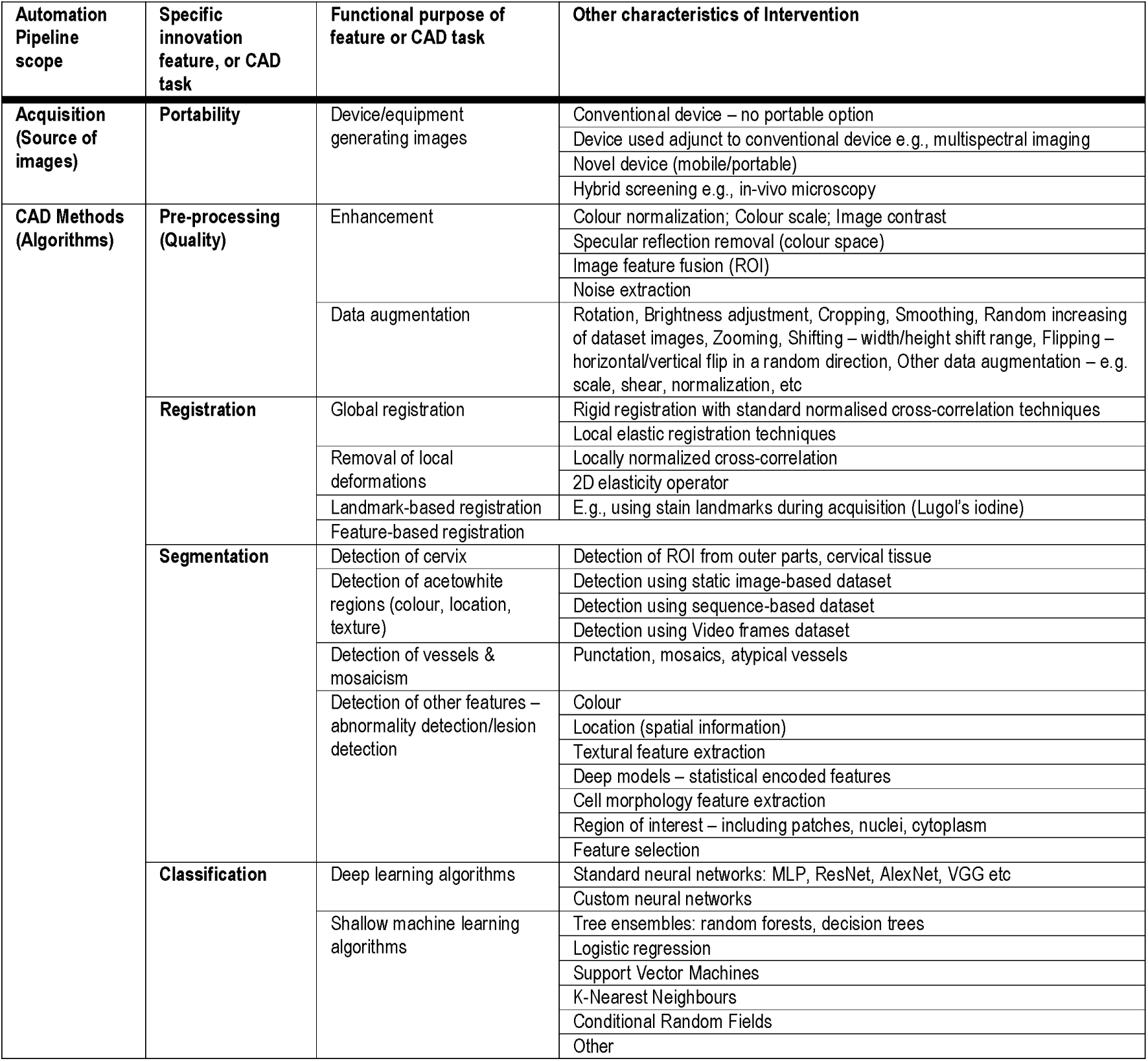
Integrated overview of the full automation pipeline.

The CAD pipelines in all three cervical cancer screening domains can all be described the same way, as indicated by the themes grouped as *‘Specific innovation feature, or CAD tas*in*k’*Table 1. Some authors’ contributions spanned the entire automation pipeline from acquisition to classification (n=17), while other contributions covered a portion of the pipeline via standalone algorithms (n=56).

Variations in contributions became evident when looking at functional purpose and other characteristics of interventions. This was not surprising because the design of an algorithm depends on the properties of the images being analysed, which vary considerably between domains.

Findings under each innovation feature (or CAD task) in Table 1 are summarized as follows:

#### Acquisition: Portability

Portable screening devices were identified from 17 studies, and these are listed in Supplementary Table 3. The table contains a summary of the portable devices for each screening domain, and a summary of the stages of the medical device development cycle where the devices lie. The stages were synthesised based on three stages defined by the medical device development lifecycle framework, viz concept & feasibility phase, verification and validation phase (pre-launch clinical trials are conducted in this phase); and c) product launch & post market monitoring phase i.e. the phase of clinical adoption [33]. Ten studies used CAD-enabled portable devices in the feasibility phase [34,35, 36,37,38,39,40,41,17,42]. Six studies used CAD-enabled portable devices which were in the verification and validation phase [32,18; 43,44,45,16]. Of the six studies, three studies used the same device (MobileODT EVA) but applied different algorithms. One study used a CAD-enabled portable device, i.e., the Grandium Ocus which was in the product launch & post monitoring phase at the time of writing [46].

Portable micro-endoscopy systems were identified, these were the mHRME [37] and the PiHRME [41]. These systems are an extension of mobile colposcopy; however, visualization of abnormality is conducted at subcellular level [22]. Thus, these portable microscopes can be considered to be in a hybrid screening domain since their application lies at the intersection of colposcopy and cytology (or microscopy).

Commercial cytology scanners that were identified were not portable, these were the BD FocalPoint™ GS imaging system, and the Hologic ThinPrep® Imaging system [47]. Two additional devices were identified however it is unclear whether they are commercially available; these are the high-resolution slide scanner iScan 2.0 [48] and the signal plane scanner [49].

Novel histopathology systems were used adjunct to conventional microscopes; therefore, they were not fully portable. Only the supplementary parts being paired to conventional devices were portable. The systems identified were the fluorescence lifetime imaging system [50], the confocal fluorescence imaging system [51], and the snapshot narrow band imaging camera [52]. Confocal fluorescence acquisition can be performed in-vivo. This is advantageous since histopathological results can be obtained in real-time during a patient visit instead of patients waiting for weeks for results from pathology laboratories [51]. Furthermore, the possibility of in-vivo pathology could reduce the costs associated with biopsy excision and analysis in the laboratory.

#### Pre-processing

Depending on the image acquisition device used within a cervical screening domain, different image quality issues are encountered [53]. Common pre-processing techniques in colposcopy were colour space subtraction, adaptive thresholding, filling, and filters. These techniques were used to achieve automated speculum removal [54, 36, 18, 55]. The majority of studies using cytology images used techniques for denoising and tissue contrast enhancement [56; 50; 57,58]. An additional common pre-processing task across all screening domains was data augmentation, which is a technique used to avoid model overfitting when training deep learning models with low volumes of data.

#### Registration

Registration was used more frequently in the colposcopy domain, e.g. rigid global registration [59], registration on time-series images [60], and landmark-based registration [61,42,40]. In the cytology and histopathology domains, feature-based registration [62] and spatial co-registration of images [52] were used.

#### Segmentation and classification

From the relevant reviewed studies, one of two machine learning approaches (shallow or deep learning) were observed for segmentation and classification algorithms. Table 2 lists the studies where segmentation and classification were applied. Summaries of observations from each screening domain are presented below.

**Table 2:** Segmentation and Classification CAD outcomes.

| Outcome | Colposcopy studies | Cytology studies | Histopathology studies | Microendoscopy studies |
| --- | --- | --- | --- | --- |
| **Segmentation (ROI detection outcome)** | Device linked to algorithm:<br>None<br>Standalone CAD (n=7):<br>Guo et al. (2020) [Mask R-CNN]<br>Cholkeri-Singh et al. (2018) [DSI feature map]<br>Shrivastav et al. (2018) [canny edge detection model & Earth Movers Distance]<br>Yan et al. (2022) [HLDnet]<br>Bai et al. (2018) [K-means clustering algorithm]<br>Srinivasan et al. (2009) | Device linked to algorithm (n=1):<br>Dalla Palma et al. (2013)[CAD unspecified]<br>Standalone CAD (n=6):<br>Wang et al. (2021) [FCN]<br>Li et al. (2021) [DGCA-RCNN]<br>Ma et al. (2020) [FPN]<br>Qin et al. (2015)[super pixel-based segmentation]<br>Muhimmah et al. (2017)[Watershed transformation] | None | None |
|  | [GMM]<br>Das et al. (2011)[K-means clustering algorithm] |  |  |  |
| **Binary classification outcome** | Device linked to algorithm(n=9):<br>Asiedu et al. (2018) [SVM classifier]<br>Park et al. (2021)[XGB,RF, SVM, ResNet-50]<br>Huh et al. (2004) [multivariate statistical analysis]<br>Monsur et al. (2017) [mean gray values]<br>Kudva et al. (2018a) [SVM]<br>Bae et al. (2020) [KNN]<br>Vinals et al. (2021) [ANN]<br>Peterson et al. (2016) [CervDx Application]<br>Kudva et al. (2018b) [shallow CNN]<br>Standalone CAD (n=11):<br>Gutiérrez-Fragoso et al. (2017) [KNN]<br>Mary et al. (2012) [CRF]<br>Hu et al. (2018) [Faster R-CNN]<br>Li et al. (2020) [E-GCN]<br>Kudva et al. (2020) [Custom CNN]<br>Park et al. (2011) [CRF]<br>Xue et al. (2020b) [CAIADS: YOLO, U-Net]<br>Peng et al. (2021) [VGG16]<br>Saini et al. (2020) [ColpoNet]<br>Mehlhorn et al. (2012) [KNN]<br>Jaya et al. (2018) [ANFIS classifier] | Device linked to algorithm(n=3):<br>Wang et al. (2009) [FISH]<br>Yamal et al. (2015)[DT]<br>Holmström et al. (2021)[CNN]<br>Standalone CAD (n=7):<br>Bhowmik et al. (2018) [SVM]<br>Gertych et al. (2012) [SVM]<br>Zhang et al. (2014) [RF]<br>Ke et al. (2021) [ResNet-50]<br>Sornapudi et al. (2019)[VGG19 and ResNet-50]<br>Sun et al. (2017) [RF]<br>Schilling et al. (2007)[FLD] | Device linked to algorithm(n=1):<br>Pal et al. (2021) [Deep Multiple instance learning algorithm]<br>Standalone CAD algorithms (n=5):<br>Gu et al. (2015) [Extreme learning machine]<br>Yi et al. (2020) [Euclidean distance algorithm]<br>Rahmadwati et al. (2012) [Hybrid of graph cut and colour segmentation]<br>Konstantinou et al. (2018) [Wilcoxon statistical test and Biserial correlation]<br>Zhao et al. (2016)[SVM] | Device linked to algorithm (n=2):<br>Hunt et al. (2018) [un disclosed CAD algorithm]<br>Parra et al. (2020) [MobileNetV2]<br>Standalone CAD: None |
| **Multiclass classification outcome** | Device linked to algorithm(n=1):<br>Chandran et al. (2021) [CYENET]<br>Standalone CAD (n=4):<br>Yuan et al. (2020) [ResNet-50, Mask R-CNN]<br>Chen et al. (2019) [RF]<br>Buiu et al. (2020) [MobileNet V2]<br>Novitasari et al. (2020) | Device linked to algorithm(n=1):<br>Sampaio et al. (2021) [F-RCNN]<br>Standalone CAD (n=13):<br>Sokouti et al. (2014) [LMFFNN]<br>Hussain et al. (2020) [CNN Ensemble]<br>Bhatt et al. (2021) [EffecientNet-B3]<br>Lin et al. (2021)[rule-based risk] | Standalone CAD algorithms(n=2):<br>Sheikhzadeh et al. (2015) [Statistical correlation]<br>Meng et al. (2021) [CNNs – DeepLab v3+] | None |
|  | [KELM] | stratification (RRS) module;<br>DCNN<br>Manna et al. (2021) [ensemble of<br>CNN classifiers]<br>Shanthi et al. (2021) [SVM]<br>William et al. (2019) [Fuzzy C-means]<br>Win et al. (2020) [tree ensemble<br>classifier]<br>Zhu et al. (2021) [CNN ensemble]<br>Kim et al. (2015) [Hough<br>transform extraction algorithm]<br>Liang et al. (2021) [Faster R-CNN<br>with FPN]<br>Tripathi et al. (2021) [Resnet152]<br>Suguna et al. (2021) [GBT, NNM] |  |  |
| **Acronyms:** Mask R-CNN: Mask Region-based Convolutional Network. DSI feature map: Dynamic Spectral Imaging. HLDnet: HSIL+ Detection Network. GMM: Gaussian mixture modelling. SVM: Support Vector Machine. ResNet-50: Residual Neural Network with 50 layers. XGB: X Gradient Boost. KNN: K-Nearest Neighbours. CRF: Conditional Random Forests. ANN: Artificial neural network. RF: Random Forests. YOLO: You Look Only Once. E-GCN: Graph Convolutional Network with Edge features. CAIADS: Colposcopic Artificial Intelligence Auxiliary Diagnostic System. CYENET: Colposcopy Ensemble Network. CNN: Convolutional Neural Network. KELM: Kernel Extreme Learning Machine. VGG-16: Visual Geometry Group with 16 layers. ANFIS: Adaptive Neuro Fuzzy Inference System classifier. FCN: Fully Convolutional Network. DGCA-RCNN: Faster Region-based Convolutional Neural Networks with infused deformable and global context aware layers. FPN: Feature Pyramid Network. FISH: Fluorescent In-Situ Hybridization. DT: Decision Trees. ResNet-152: Residual Neural Network with 152 layers. VGG-19: Visual Geometry Group with 19 layers. LMFFNN: Levenberg-Marquardt feedforward MLP neural network. DCNN: data-driven deep convolutional neural network. FLD: Fischer Linear Discriminant. SGF: Statistical Geometric Features. GBT: Gradient Boosting Tree. NNM: Neural network model.<br>Notes:<br>1. Deep learning algorithms are highlighted in grey<br>2. The type of algorithm used by Peterson et al. (2016) [CervDx Application] cannot be determined from the given name of the model.<br>3. The type of algorithm used by Dalla Palma et al. (2013) is not known. |  |  |  |  |

##### Colposcopy segmentation and classification

Within the reviewed literature, algorithms detecting acetowhite regions used static and/or temporal image datasets (i.e., sequence-based images or stills from video footage). Both steps were performed using algorithms which extracted features related to colour information, texture information or spatial information [63, 54; 18, 45]. Colour features were largely used for detection of cervix and acetowhite regions. Whereas texture features were largely used for the detection of atypical vessels and mosaicism. It was observed that the majority of colposcopy studies (18 out of 32) which were reviewed used shallow machine learning algorithms for CAD segmentation and classification.

##### Cytology segmentation and classification

For the majority of reviewed cytology studies, image segmentation involved detection of ROIs from whole slide image datasets (n=17). Three studies used single cell datasets [35,64, 58]. Feature extraction algorithms were used to identify and select features related to colour information, texture information, and cell morphology (that is, area and shape of nuclei, and area of cytoplasm).

It was observed that a slight majority of reviewed cytology studies (15 out of 30) used deep learning algorithms for CAD segmentation and classification. In contrast, 14 out of 30 cytology studies used shallow machine learning algorithms.

##### Histopathology segmentation and classification

Histopathology classification algorithms identified in the review used cell morphology and textural features to identify abnormal ROIs. It was observed that the majority of reviewed histopathology studies (5 out of 8), used shallow machine learning algorithms for CAD segmentation and classification.

##### Micro-endoscopy segmentation and classification

One of the micro-endoscopy binary classification algorithms that was used was a deep-learning algorithm [41]. The algorithm used in the second micro-endoscopy study for binary classification was not disclosed [37].

### Comparator analysis

A classifier algorithm’s performance is dependent on the ground truth comparator used, and comparators reported by studies were diverse. Common performance measures reported by reviewed studies were accuracy, sensitivity (also known as recall), specificity, and area under curve (AUC). Some studies reported two or three performance measures while others reported four metrics. Deep learning algorithms used in cytology have more frequently reported greater performance measures when compared to the shallow algorithms. The comparative analysis of results for all studies that reported segmentation or classification results are summarized in Table 3.

**Table 3:**
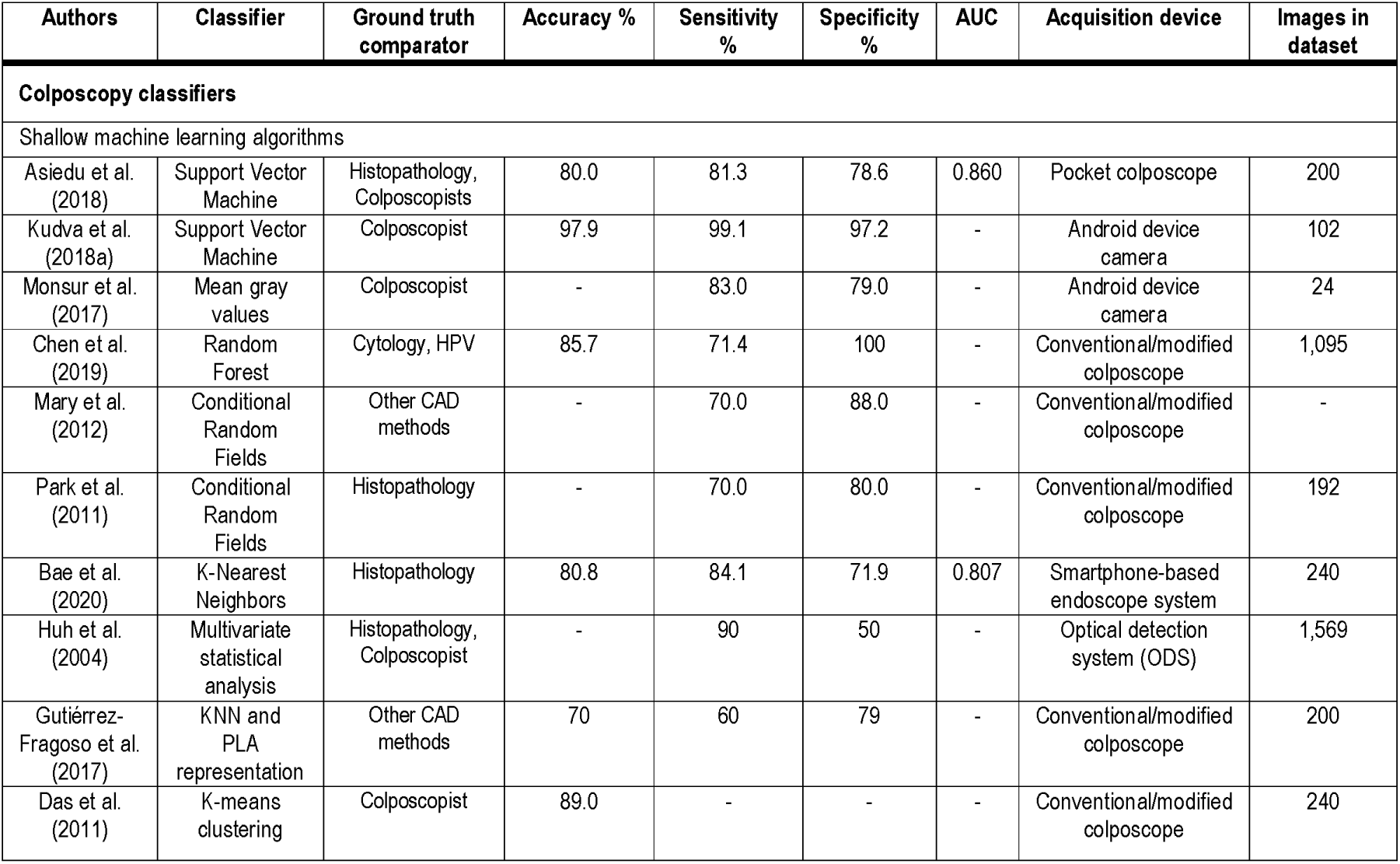
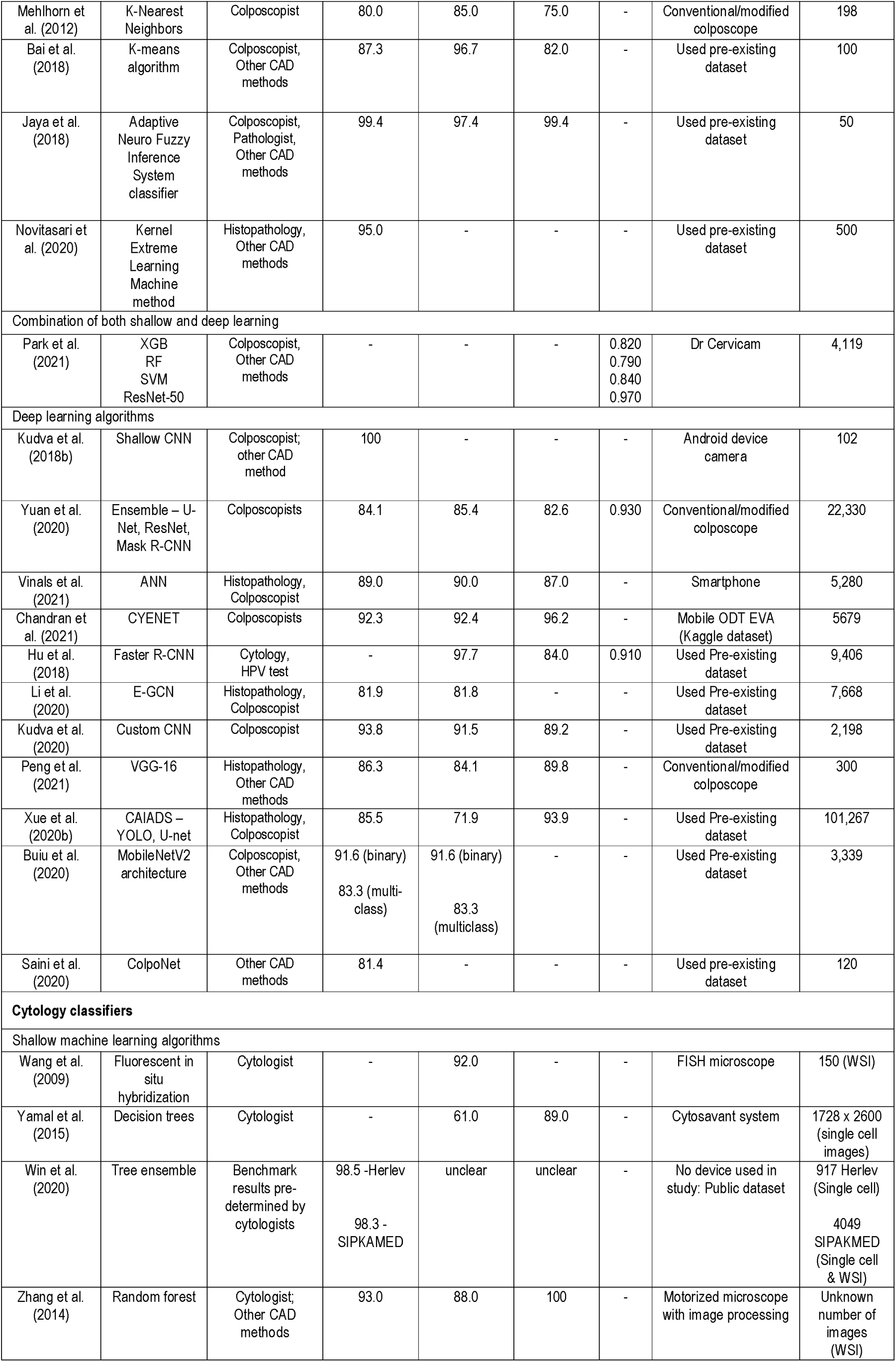
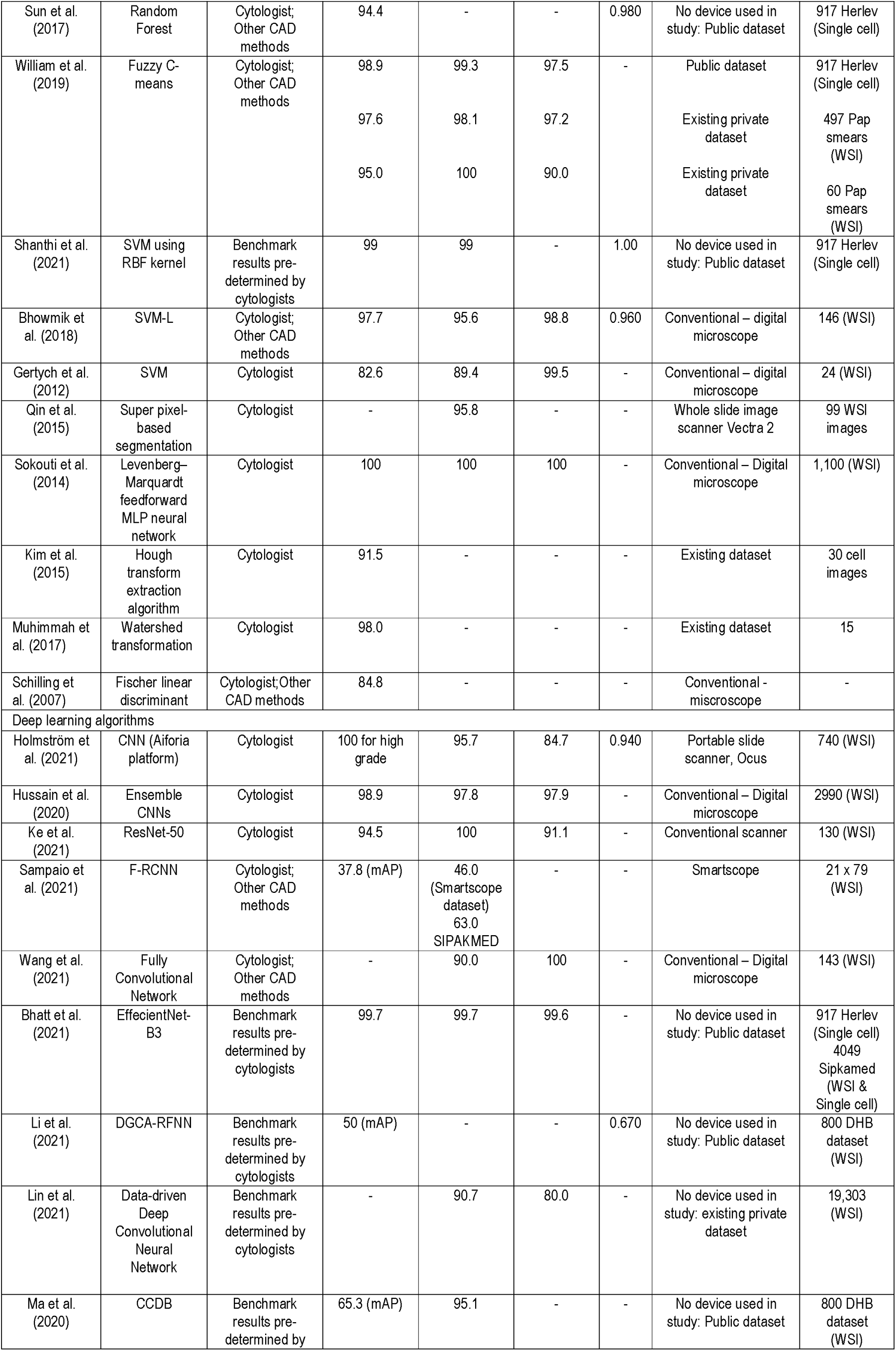
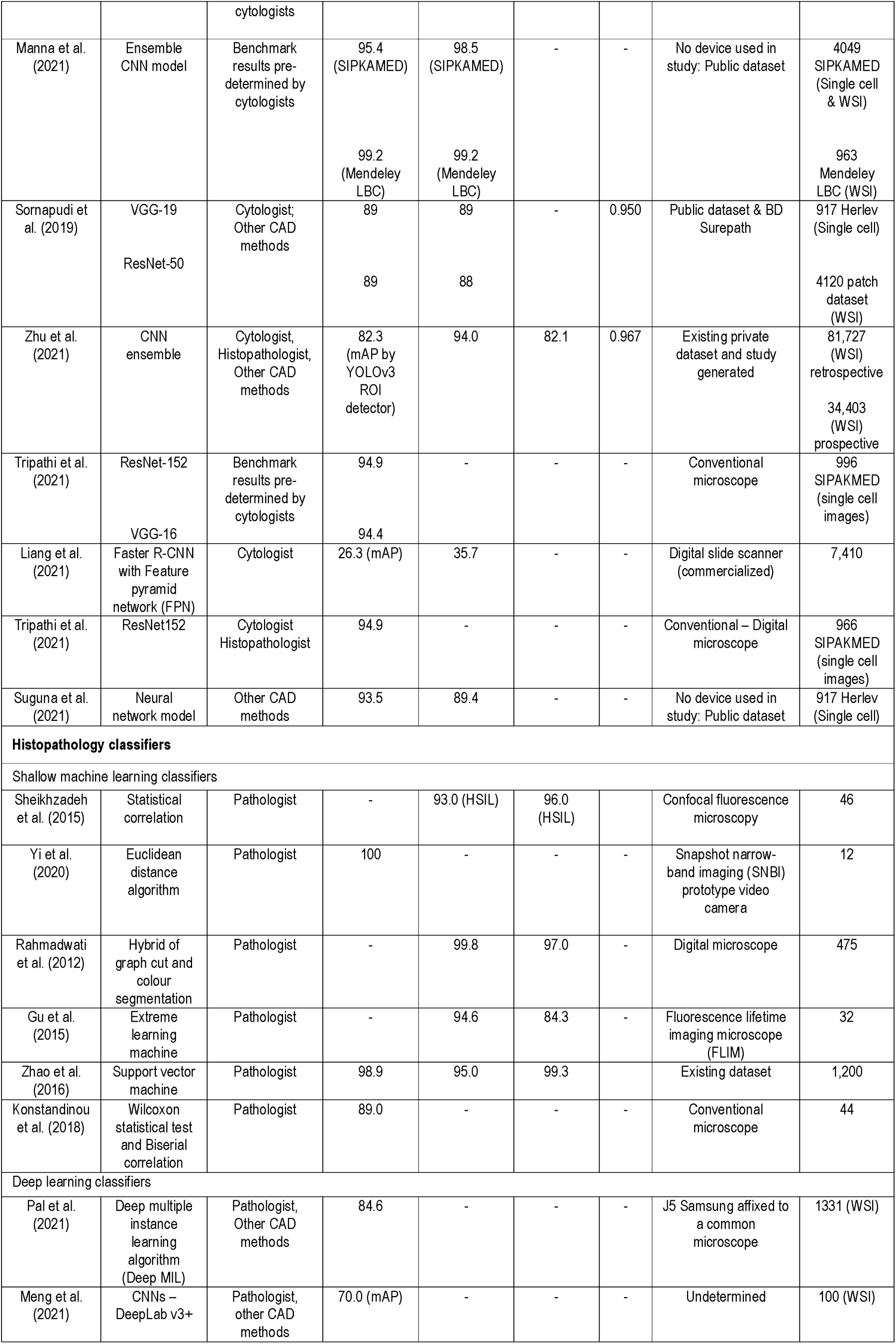
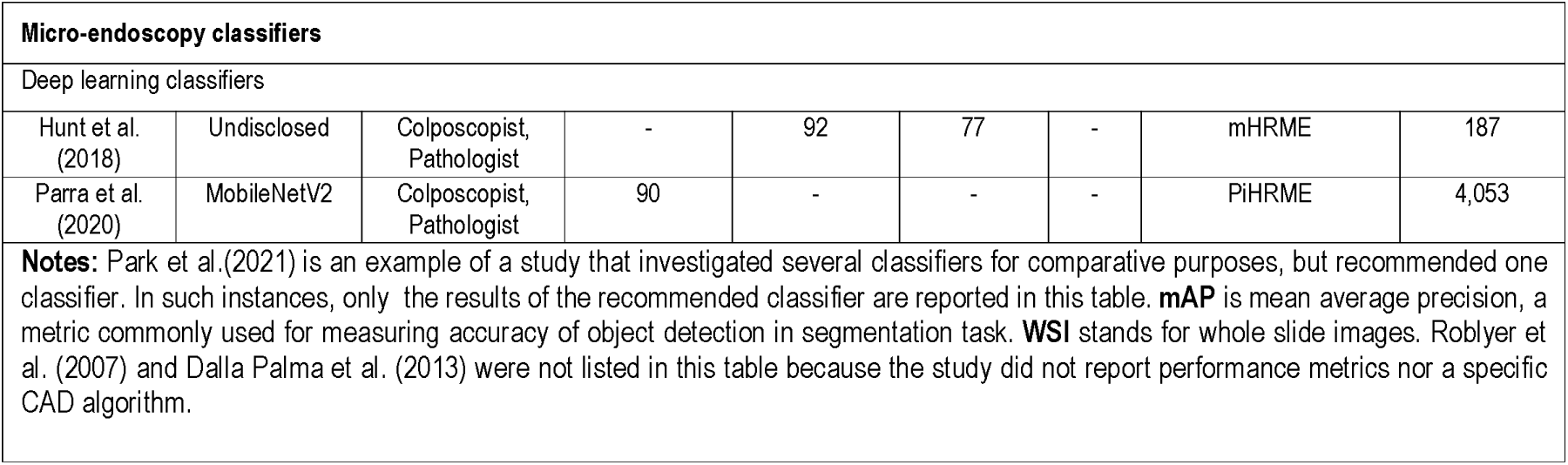
Comparison of state-of-the-art classifiers reported in literature.

### Outcome analysis

For segmentation, the study outcomes were ROI detection. For classification, the study outcomes were binary or multiclass classification. Seventy-two studies reported CAD outcomes and only one study did not report the outcome of automated analysis [65]. Seventeen studies used CAD-enabled devices and 53 studies used standalone algorithms to determine decision outcomes. As seen in Table 2, a total of 13 studies only reached the segmentation step by yielding ROI detection outcome, and a total of 59 studies fulfilled the last step of the automation pipeline by achieving a classification outcome.

### Quality assessment

This systematic review found that the majority of studies had low risk of bias. However, this review has high publication bias in general due to lack of grey literature with adequate information to evaluate PICO criteria. The methodological bias in some studies was due to issues of ethical approval, unclear steps in the method, unknown accuracy of results and/or no disclosures of conflicts of interest. Regardless of the high risk of methodological bias, the studies were included in analyses because they met PICO criteria and their qualitative findings were relevant to this systematic review.

## 3. Discussion

### General discussion on interventions

Two types of interventions were found, i.e., 1) integrated systems which had an acquisition device linked to an image analysis algorithm; and 2) standalone image analysis algorithms. Seventeen studies used acquisition devices with corresponding image analysis algorithms. These device and algorithm systems are presented in Supplementary Table 3. Most of these devices were designed using smartphones; they include the Pocket Colposcope, MobileODT EVA Colpo, Smartphone Camera, Smartphone-based Endoscope System, Smartscope, mHRME and PiHRME. The majority of included studies reported standalone image analysis algorithms (Table 2 shows groupings of standalone algorithms in each screening domain).

### Fundamental goals of interventions

The contributions of studies were considered valuable if they promoted any of the following fundamental goals: 1) obtaining clinical validation within LMIC contexts; 2) having automated systems achieve greater efficacies than manual analysis; and/or 3) lowering the cost of cervical cancer screening program(s). Interventions consisting of acquisition and automated analysis met all the fundamental goals. In contrast, standalone algorithms achieved only one goal (greater prediction performance), and without pairing them with an inexpensive portable acquisition device, the standalone algorithms would not be suitable in LMIC.

### Goal 1(a) of clinical validation in LMIC as it relates to population

According to the Food & Drug Administration (FDA), true clinical validation is achieved when the intended purpose of an intervention is proven reliably within the target population [68]. It has not been proven whether diagnostic performances attained with current interventions could be sustained if the study methodology were designed for nurses as crucial users of automated interventions, instead of specialist clinicians.

### Goal 1(b) of clinical validation in LMIC as it relates to the intervention dimension of PICO

The CAD models associated with mobile acquisition devices in colposcopy tend to be shallow machine learning algorithms [18,38], because shallow models have faster computational speeds and robust performance on small datasets [45]. The limited battery life of mobile phones precludes the use of energy-intensive neural network-based algorithms, especially when there is no guarantee of readily available battery charging solutions [16].

Cloud computing has been suggested as a possible solution to accommodate energy-intensive applications. However, the implementation of cloud computing needs to be investigated in LMICs because cloud-based systems need good network connectivity. Peterson [32] reported that having good network connectivity was not always feasible at a screening camp in Kenya. Roblyer et al. [65] reported challenges when using their device in a pre-trial feasibility study conducted in Nigeria. One of the challenges was unstable electricity supply, which is a frequent occurrence in many LMICs. MobileODT, has adapted their commercialised AVE solution, which is a cloud-based web service, to run on the EVA Colpo smartphone device via a mobile application (MobileODT, 2019). Currently, clinical validation of the adapted algorithm and mobile application are underway in LMICs globally [69,70].

The disadvantage of shallow machine learning algorithms is that they underperform against deep learning alternatives mostly when applied on multi-classification problems. This implies that there is a trade-off that researchers have to make between computational simplicity and prediction accuracy. Research is ongoing on adaptations of deep learning models that are necessary for the models to be embedded onto mobile apps [66]. In addition, research is growing on edge computing, where edge platforms can operate independently during network outages [71]. Sun [72] reported a prototype Google Edge TPU which, when embedded to cameras, could allow computations of neural networks.

A common path of development has been observed, where the first component, the device, is clinically validated and commercialized; then the second component, the algorithm, subsequently goes through its own validation track. Eventually, the two validated modules are merged to provide an end-to-end product. However, no studies in colposcopy have reported reaching the point of fully validating and commercializing a combined system of mobile device and app-based analysis, regardless of that being indicated as the desired end point that researchers are aiming for. Of the portable colposcopy technologies reviewed, MobileODT seems to be the only example whose clinical validation in LMIC is fairly advanced.

### Goal 2 on higher efficacies as it relates to the comparator dimension of PICO

The performance of automated image analysis systems is measured by any of the following common parameters: precision, accuracy, sensitivity, specificity and area under the curve (AUC). The higher these parameters are, the better the performance of the algorithm. If accuracy is used in isolation, it can be a misleading metric when a dataset has an imbalance skewed towards the negative class. In screening programs, there’s a prevalent trend of negative case skewness within datasets, attributed to the predominance of negative outcomes over positive ones in the collected data. [74]. Hence, metrics like sensitivity and specificity succinctly enhance performance analysis in skewed datasets. Area under curve is usually the best way to compare various binary classifiers’ performances because an AUC-maximising classifier has the advantages of robust performance even under data imbalanced conditions. Any binary classifier yielding the greatest AUC would be the model proven superior over others. However, such a comparison was difficult to make in this systematic review because there was inconsistency in reporting of AUC, with some studies reporting it and some not. This made selection of the best system on the basis of accuracies challenging.

#### Colposcopy prediction performance

Studies on automated image analysis in colposcopy indicated accuracies ranging from 70.0% – 100%; sensitivities ranging from 81.3% – 98.1%; and specificities ranging from 50.0% – 98.5%. In comparison, the accuracy range of manual colposcopy analysis, according to Asiedu et al. [18], was typically 52.0% – 70.0%; sensitivities typically ranged from 68.0% – 88.0%; and specificities typically ranged from 20.0% – 71.0%; provided histopathology was the ground truth against specialist judgement. This provides evidence that automated interventions in colposcopy, produce more effective diagnostic results compared to results attained through manual diagnosis. However, the upper limits of the performance ranges from automated analysis were skewed by presence of studies utilising subjective colposcopy judgement only, as ground truth. These types of studies more frequently achieved performance values above 90.0%. In contrast to this, studies that used histopathology as ground truth, most frequently had performance values under 90.0%. If the ranges were evaluated with only the subset of studies that indicated histopathology as ground truth, then sensitivity estimates for automated systems in the subset would range from 80.0% – 90.0%. Since this subset of sensitivity estimates vary marginally from sensitivities of manual analysis (claimed to range between 68% – 88%), it was not always true that automated analysis yielded superior sensitivities than manual analysis. On the other hand, it can indeed be said that automated interventions unconditionally yielded superior accuracy and specificity ranges. Even when a subset of pathologically confirmed studies was considered, accuracies from automated analysis remained greater, ranging from 80.0%-99.4%, and specificities also remained greater, ranging from 79.0%-93.0%. These deductions point to a need for a meta-analyses to conclusively determine whether automated analysis yields greater prediction performance than manual analysis.

#### Cytology prediction performance

According to Kanavati et al. [75] commercial automated cytology scanners did not produce greater performance metrics for all classes (particularly the class of atypical squamous cells of undetermined significance) than conventional manual analysis, but rather produced equivalent performance. This contradicts the observation that automated cytology scanners improve prediction performance [76]. This difference in views might have stemmed from there being marginal differences between automated analysis and manual analysis, as was the case for colposcopy. In cytology, research seems to have focussed on improving automation model performance through application of deep learning algorithms, which are more robust for multiclass abnormality classification than shallow machine learning algorithms. However, as indicated in the previous sections, shallow machine learning algorithms of similar performance level may be advantageous over deep models due to computational simplicity, which may allow porting to a smartphone [66]. Prediction performance is an important success factor that researchers seem to have focused on. However, since performance alone is not enough to justify any decision for implementation of an automated system in LMICs, other factors for evaluating successes of an automated screening system should be equally considered. That is, factors such as cost of intervention, and degree of clinical validations by developers or researchers. These are further described in the following paragraphs.

### Goal 3 on lower screening program cost

Screening program expenses consist mainly of infrastructure/equipment and labour costs. Equipment costs in particular can be reduced by utilizing inexpensive devices. However, most studies presenting novel alternatives did not report on costs. Since most portable acquisition devices in colposcopy were designed for integration with smartphones, the expectation of cost reduction is based on the assumption that the purchase cost of mobile phones is more affordable than conventional medical imaging devices. However, purchase price alone is not a good reflection of the total costs of a device if hidden costs have been disregarded. Purchase price makes up to 20-30% of total cost of a product during its lifecycle, and hidden costs make up to 70 – 80% of the full costs of a product. Examples of hidden costs are operational costs such as consumables or parts, maintenance/upgrade costs, training and education, downtime, et cetera [77]. Researchers have not assessed how the total lifecycle costs of novel devices compare against the lifecycle costs of conventional devices. In cytology and histopathology, it was not demonstrated whether the purchase prices of novel devices that were proposed were low-cost when compared to conventional microscopes or scanners.

Health technology assessments (HTA) are an option that researchers can use to address unknowns about the cost-effectiveness of technologies. This type of research study is often needed for justifying policy decision on adopting a medical technology [47,78]. From the analysed studies, only commercial automated technologies in cytology have been investigated by a health ministry for adoption into national standards through an HTA study [47]. Since national healthcare standards developed by policy makers greatly influence widespread adoption in clinical practice [78], any technology with favourable outcomes from an HTA study would most likely gain greater success.

### Recommendations

#### Meta-analysis

The qualitative analysis of prediction performance supported that automated systems generate greater efficacies. However, because some parameters were marginally higher than manual analysis, a meta-analysis will be required in the future to statistically prove the hypothesis that automated analysis yields greater efficacies than manual analysis. Such a meta-analysis would require researchers to standardise performance metrics.

#### Economic analyses

Since most portable colposcopy acquisition devices were developed using smartphones, any cost savings assume that mobile phones are cheaper than other medical imaging devices. However, this assumption must be interrogated because the purchase price alone indicates a small fraction of the total costs of the product over its lifetime. Costs such as internet data, cloud computing fees, and costs of biennial device upgrades (as hardware needs to keep up with software upgrades) are some examples of hidden costs that need to be considered to determine the true cost-effectiveness. It is unclear what the economic feasibility of proposed interventions will be over a lifetime period compared to that of a conventional device.

#### Clinical validations

It is unknown if the diagnostic performance attained with current automated interventions could be maintained when nurses use the automated interventions instead of expert physicians.

According to Kumar et al. [67], while AI-based models have demonstrated their superiority in cancer research, their practical implementation in low-resource clinics is lacking. This is possibly due to inadequate data availability since many LMIC do not have electronic medical records, nor national digital health strategies; there is poor electricity supply or internet infrastructure to support implementation of digital solutions [79]. This systematic review revealed one area for improvement in clinical validations; that researchers must evaluate a fully integrated system rather than only testing components of a system in isolation. Evaluation of the entire system during both short and long equipment run times is the only way to eliminate assumptions thereby providing a fact basis on the resilience of a system as a whole, the true computation performance, and prediction of performance results [80]. Many proposed models are yet to be comprehensively validated in clinical LMIC settings to ensure that implementation challenges unique to low resource settings are addressed by proposed systems.

## 4. Conclusion

Seventeen studies reported the application of algorithms coupled to novel image acquisition devices, and 53 studies reported standalone image analysis algorithms. The main advantage of the novel acquisition devices identified was that they were based on mobile phone technology and could therefore have low purchase costs, unlike conventional alternatives which are heavy and expensive. Apart from portable/mobile CAD-enabled devices, other innovative CAD-enabled device(s) were identified, for example, a device that was used adjunctive to a microscope in histopathology. This device is the confocal fluorescence acquisition system, and it overlaps two cervical cancer domains, namely colposcopy and histopathology because it can visualize tissue in-vivo for high-grade CIN detection. This means this technology could have an impact of optimizing the cervical cancer screening process; thus, potentially giving patients the convenience of receiving histopathological results the same day of their visit, instead of waiting weeks for results to come back from a central laboratory.

Automated analysis improved diagnostic decisions made by clinicians by achieving higher prediction accuracies than manual expert analysis. The type of CAD algorithms identified were either deep machine learning algorithms or shallow machine learning algorithms, and they were used to detect ROIs in an image, or to classify normal cases from abnormal cases. Deep learning methods, which are more reliable for classifying multiclass abnormalities than shallow machine learning algorithms, have been used more commonly in cytology research to improve automation model performance especially for granular multiclass classification problems. However, alternative shallow machine learning algorithms were reported to be advantageous over deep models, due to computational simplicity, which is considered to be more suitable for execution on mobile phones. This implies that there is a trade-off that researchers make between computational simplicity and prediction accuracy. Although the majority of studies focused on standalone algorithms, with the goal of improving prediction performance, performance alone is insufficient to justify the implementation of an automated system in LMICs. Other aspects, such as intervention costs and stage of development, should be considered when evaluating the feasibility of an automated cervical cancer screening system.

Research groups tend to focus on their respective disciplinary areas, for example, medical device development or software development, before cross-collaborating to commercialize integrated technology systems. In colposcopy, no developer had completed the validation of an integrated device and software system. However, one developer (i.e. MobileODT) was more advanced since their clinical validations for an integrated system was in progress at the time of this systematic review. When conducting clinical validations in LMIC, it is crucial to use algorithms on mobile devices to minimise reliance on connectivity to cloud platforms. Furthermore, it is necessary to validate if performance of interventions is maintained when the intervention is applied by nurses instead of specialists. This paper will hopefully draw more attention to automated systems featured herein, so that the promising systems may be advanced into clinical trials and health technology assessments in LMICs.

## Data availability

No new data were generated or analysed for the manuscript.

## Acknowledgements

The Groote Schuur Hospital Obstetrics and Gynaecology team running the Khayelitsha Cervical Cancer Screening Project, provided learning opportunities through practical exposure to the real-world application of one on the automated devices developed by MobileODT. We thank Professor Lynette Denny for encouragement and support.

## Author Contribution Statement

LL, RS, LK, BM and TM defined the research topic. LL registered the protocol, conducted and wrote the systematic review. BM, RS and TM supervised the systematic review. RS screened articles. TM mediated conflicting decisions on screened articles. LL, BM, RS, LK and TM made revisions to the article. All authors read and approved the final manuscript.

## Research Contribution

This systematic review provides a perspective on CAD-enabled cervical cancer screening technology systems for the cytology, colposcopy, and histopathology domains of cervical cancer screening protocols. In contrast to our work, the majority of prior reviews examined only a single domain. This is a fragmented approach, in our view, because successful diagnosis depends on all three domains, which have common challenges of specialist shortages and expensive equipment. The similar challenges across multiple domains provides an opportunity to discover unforeseen overlapping technologies. In this study, the multi-domain methodology helped identify two novel technologies that overlap across two domains, which might have otherwise been missed. The two overlapping technologies are the mobile high-resolution micro-endoscopy (mHRME) and confocal fluorescence imaging. High resolution micro-endoscopy intersects between colposcopy and cytology domains, and confocal fluorescence imaging intersects between colposcopy and histopathology domains. In addition, this systematic review identifies pre-commercial and commercialized technologies used across the full automation pipeline which consists of acquisition, pre-processing, registration, segmentation and classification. Including image acquisition in the automation pipeline is necessary because the types of CAD algorithms that may be used for the subsequent automated image analysis are determined by the image acquisition device used.

## Scope and limitations

The reader should note the following scope: This systematic review focused on interventions in cervical cancer screening, and not staging. Moreover, relevant CAD-enabled cervical cancer screening technology systems were those producing optical images with decision-support, which were applied in the cytology, colposcopy, and histopathology domains of the cervical cancer screening protocol. Particular attention was given to interventions in these screening domains because portable devices could be used to produce optical images.

