## Supplemental Tables for "Automated analysis of digital medical images in cervical cancer screening: A systematic review"

**Supplementary Tables and Figures**

Supplementary Table 1: PubMed Search strategy

| **No.** | **Description** | **Search Query** | **# of hits** |
| --- | --- | --- | --- |
| #1 | Free text | (Women) OR (low resource countries) OR (resource-limited countries) OR (low- middle-income countries) OR (developing countries) OR (rural areas) | 1,694,734 |
| #2 | Mesh terms and free text | ("Uterine Cervical Neoplasms"[Mesh]) OR (cancer cervix)) OR (gynaecological cancer)) OR (cervical cancer) | 221,415 |
| #3 | Mesh terms | ("Diagnosis"[Mesh] OR "Diagnostic Screening Programs"[Mesh] OR "Early Detection of Cancer"[Mesh]) OR "Mass Screening"[Mesh] | 8,982,055 |
| #4 | Mesh terms and free text | ("Image Processing, Computer-Assisted/classification"[Mesh]) OR ("Image Processing, Computer-Assisted"[Mesh]) OR (computer-aided analysis) OR (automated image analysis) OR ("Pattern Recognition, Automated"[Mesh]) OR ("Telemedicine/classification"[Mesh]) OR ("Mobile Health Units"[Mesh]) OR (mHealth) OR (digital health) OR (portable digital image devices) OR (digital alternative techniques for detection of CIN) OR (digital colposcopy) OR (digital cytology) OR (digital microendoscopy) OR (digital cervical histopathology) OR (digital multispectral imaging of cervix) | 393,051 |
| #5 | Mesh terms and free text | ("Papanicolaou Test/classification"[Mesh]) OR ("Cytological Techniques"[Mesh])) OR ("Human papillomavirus 16"[Mesh])) OR ("Colposcopy/classification"[Mesh])) OR ("Image-Guided Biopsy"[Mesh])) OR (colposcopy-directed biopsies)) OR ("Anatomic Landmarks/diagnostic imaging"[Mesh])) OR (visual inspection lugol’s iodine)) OR (VILI)) OR (visual inspection with acetic acid)) OR (VIA)) OR (VIAM)) OR (microendoscopy) | 2,843,625 |
| #6 | Mesh terms and free text | (Automated diagnosis) OR (automated diagnostic decision)) OR (automated detection)) OR (decision support) | 384,920 |
| #7 | #1 AND #2 AND #3 AND #4 AND #5 AND #6 | (Women) OR (low resource countries) OR (resource-limited countries) OR (low- middle-income countries) OR (developing countries) OR (rural areas) AND (("Uterine Cervical Neoplasms"[Mesh]) OR (cancer cervix)) OR (gynaecological cancer) OR (cervical cancer) AND (("Diagnosis"[Mesh] OR "Diagnostic Screening Programs"[Mesh] OR "Early Detection of Cancer"[Mesh]) OR "Mass Screening"[Mesh]) AND (("Image Processing, Computer-Assisted/classification"[Mesh]) OR ("Image Processing, Computer-Assisted"[Mesh])) OR (“computer-aided analysis”) OR (“automated image analysis”) OR ("Pattern Recognition, Automated"[Mesh]) OR ("Telemedicine/classification"[Mesh]) OR ("Mobile Health Units"[Mesh]) OR (mHealth) OR (“digital health”) OR (portable digital image devices) OR (digital alternative techniques) OR (detection) OR (CIN) OR (digital colposcopy) OR (digital cytology) OR (digital microendoscopy) OR (digital cervical histopathology) OR (digital multispectral imaging) AND (("Papanicolaou Test/classification"[Mesh]) OR ("Cytological Techniques"[Mesh])) OR ("Human papillomavirus 16"[Mesh]) OR ("Colposcopy/classification"[Mesh]) OR ("Image-Guided Biopsy"[Mesh]) OR (“colposcopy-directed biopsies”) OR ("Anatomic Landmarks/diagnostic imaging"[Mesh]) OR (“visual inspection”) OR (VILI) OR (“lugol’s iodine” ) OR (“acetic acid”) OR (VIA) OR (VIAM) OR (microendoscopy) AND ((“Automated diagnosis”) OR (automated diagnostic decision)) OR (“automated detection”) OR (“decision support”) | 45,108 |
| #8 | Free text, Meshterm, Text Word | ((((Women) OR (low resource countries) OR (resource-limited countries) OR (low- middle-income countries) OR (developing countries) OR (rural areas)) AND (("Uterine Cervical Neoplasms"[Mesh]) OR (cancer cervix) OR (gynaecological cancer) OR (cervical cancer))) AND ("Diagnosis"[Mesh] OR "Diagnostic Screening Programs"[Mesh] OR "Early Detection of Cancer"[Mesh] OR "Mass Screening"[Mesh])) AND ((((((((((((computer-aided analysis[Text Word]) OR (health[Text Word])) OR (automated image analysis[Text Word])) OR (digital health[Text Word])) OR (portable digital image devices[Text Word])) OR (digital alternative techniques[Text Word])) OR (detection[Text Word])) OR (digital colposcopy[Text Word])) OR (digital cytology[Text Word])) OR (digital microendoscopy[Text Word])) OR (digital cervical histopathology[Text Word])) OR (digital multispectral imaging[Text Word])) | 13,188 |
| #9 | Titl/Abstract | ((((((((((computer-aided analysis[Title/Abstract]) OR (automated image analysis[Title/Abstract])) OR (digital health[Title/Abstract])) OR (portable digital image devices[Title/Abstract])) OR (portable digital image analysis[Title/Abstract])) OR (digital alternative techniques[Title/Abstract])) OR (digital colposcopy[Title/Abstract])) OR (digital cytology[Title/Abstract])) OR (digital microendoscopy[Title/Abstract])) OR (digital cervical histopathology[Title/Abstract])) OR (digital multispectral imaging[Title/Abstract]) | 6,518 |
| #10 | #8 and #9 | (((((Women) OR (low resource countries) OR (resource-limited countries) OR (low- middle-income countries) OR (developing countries) OR (rural areas)) AND (("Uterine Cervical Neoplasms"[Mesh]) OR (cancer cervix) OR (gynaecological cancer) OR (cervical cancer))) AND ("Diagnosis"[Mesh] OR "Diagnostic Screening Programs"[Mesh] OR "Early Detection of Cancer"[Mesh] OR "Mass Screening"[Mesh])) AND ((((((((((((computer-aided analysis[Text Word]) OR (health[Text Word])) OR (automated image analysis[Text Word])) OR (digital health[Text Word])) OR (portable digital image devices[Text Word])) OR (digital alternative techniques[Text Word])) OR (detection[Text Word])) OR (digital colposcopy[Text Word])) OR (digital cytology[Text Word])) OR (digital microendoscopy[Text Word])) OR (digital cervical histopathology[Text Word])) OR (digital multispectral imaging[Text Word]))) AND (((((((((((computer-aided analysis[Title/Abstract]) OR (automated image analysis[Title/Abstract])) OR (digital health[Title/Abstract])) OR (portable digital image devices[Title/Abstract])) OR (portable digital image analysis[Title/Abstract])) OR (digital alternative techniques[Title/Abstract])) OR (digital colposcopy[Title/Abstract])) OR (digital cytology[Title/Abstract])) OR (digital microendoscopy[Title/Abstract])) OR (digital cervical histopathology[Title/Abstract])) OR (digital multispectral imaging[Title/Abstract])) | 40 |
| #11 | Title/Abstract | ((((Automated diagnosis[Title/Abstract]) OR (automated diagnostic decision[Title/Abstract])) OR (automated detection[Title/Abstract])) OR (decision support[Title/Abstract])) OR (classification[Title/Abstract]) | 388,991 |
| #12 | #8 and #11 | (((((Women) OR (low resource countries) OR (resource-limited countries) OR (low- middle-income countries) OR (developing countries) OR (rural areas)) AND (("Uterine Cervical Neoplasms"[Mesh]) OR (cancer cervix) OR (gynaecological cancer) OR (cervical cancer))) AND ("Diagnosis"[Mesh] OR "Diagnostic Screening Programs"[Mesh] OR "Early Detection of Cancer"[Mesh] OR "Mass Screening"[Mesh])) AND ((((((((((((computer-aided analysis[Text Word]) OR (health[Text Word])) OR (automated image analysis[Text Word])) OR (digital health[Text Word])) OR (portable digital image devices[Text Word])) OR (digital alternative techniques[Text Word])) OR (detection[Text Word])) OR (digital colposcopy[Text Word])) OR (digital cytology[Text Word])) OR (digital microendoscopy[Text Word])) OR (digital cervical histopathology[Text Word])) OR (digital multispectral imaging[Text Word]))) AND (((((Automated diagnosis[Title/Abstract]) OR (automated diagnostic decision[Title/Abstract])) OR (automated detection[Title/Abstract])) OR (decision support[Title/Abstract])) OR (classification[Title/Abstract])) | 318 |

Supplementary Table 2: List of included articles

|  | **Title** | **Authors** |
| --- | --- | --- |
| 1 | Development of Algorithms for Automated Detection of Cervical Pre-Cancers with a Low-Cost, Point-of-Care, Pocket Colposcope | Asiedu, M. N. and Simhal, A. and Chaudhary, U. and Mueller, J. L. and Lam, C. T. and Schmitt, J. W. and Venegas, G. and Sapiro, G. and Ramanujam, N. |
| 2 | Quantitative screening of cervical cancers for low-resource settings: Pilot study of smartphone-based endoscopic visual inspection after acetic acid using machine learning techniques | Bae, J. K. and Roh, H. J. and You, J. S. and Kim, K. and Ahn, Y. and Askaruly, S. and Park, K. and Yang, H. and Jang, G. J. and Moon, K. H. and Jung, W. |
| 3 | Automatic segmentation of cervical region in colposcopic images using K-means | Bai, B. and Liu, P. Z. and Du, Y. Z. and Luo, Y. M. |
| 4 | Cervical cancer detection in pap smear whole slide images using convNet with transfer learning and progressive resizing | Bhatt, A. R. and Ganatra, A. and Kotecha, K. |
| 5 | Nucleus region segmentation towards cervical cancer screening using AGMC-TU Pap-smear dataset | Bhowmik, M. K. and Roy, S. D. and Nath, N. and Datta, A. |
| 6 | MobileNetV2 ensemble for cervical precancerous lesions classification | Buiu, C. and Dănăilă, V. and Răduţă, C. N. |
| 7 | Diagnosis of Cervical Cancer based on Ensemble Deep Learning Network using Colposcopy Images | Chandran, V. and Sumithra, M. G. and Karthick, A. and George, T. and Deivakani, M. and Elakkiya, B. and Subramaniam, U. and Manoharan, S. |
| 8 | An efficient cervical disease diagnosis approach using segmented images and cytology reporting | Chen, H. and Yang, L. and Li, L. and Li, M. and Chen, Z. |
| 9 | Digital Colposcopy With Dynamic Spectral Imaging for Detection of Cervical Intraepithelial Neoplasia 2+ in Low-Grade Referrals: The IMPROVE-COLPO Study | Cholkeri-Singh, A. and Lavin, P. T. and Olson, C. G. and Papagiannakis, E. and Weinberg, L. |
| 10 | [Health technology assessment report: Computer-assisted Pap test for cervical cancer screening] | Dalla Palma, P. and Moresco, L. and Giorgi Rossi, P. |
| 11 | Elimination of specular reflection and identification of ROI: The first step in automated detection of Cervical Cancer using Digital Colposcopy | Das, A. and Kar, A. and Bhattacharyya, D. |
| 12 | Automated detection of dual p16/Ki67 nuclear immunoreactivity in liquid-based Pap tests for improved cervical cancer risk stratification | Gertych, A. and Joseph, A. O. and Walts, A. E. and Bose, S. |
| 13 | Enhancement of early cervical cancer diagnosis with epithelial layer analysis of fluorescence lifetime images | Gu, J. and Fu, C. Y. and Ng, B. K. and Liu, L. B. and Lim-Tan, S. K. and Lee, C. G. |
| 14 | Cross-dataset evaluation of deep learning networks for uterine cervix segmentation | Guo P. and Xue, Z. and Rodney Long, L. and Antani, S. |
| 15 | Optimization of Classification Strategies of Acetowhite Temporal Patterns towards Improving Diagnostic Performance of Colposcopy | Gutiérrez-Fragoso, K. and Acosta-Mesa, H. G. and Cruz-Ramírez, N. and Hernández-Jiménez, R. |
| 16 | Point-of-Care Digital Cytology With Artificial Intelligence for Cervical Cancer Screening in a Resource-Limited Setting | Holmström, O. and Linder, N. and Kaingu, H. and Mbuuko, N. and Mbete, J. and Kinyua, F. and Törnquist, S. and Muinde, M. and Krogerus, L. and Lundin, M. and Diwan, V. and Lundin, J. |
| 17 | An observational study of deep learning and automated evaluation of cervical images for cancer screening | Hu, L., Bell, D., Antani, S., Xue, Z., Yu, K., Horning, M.P., Gachuhi, N., Wilson, B., Jaiswal, M.S., Befano, B. and Long, L.R |
| 18 | Optical detection of high-grade cervical intraepithelial neoplasia in vivo: results of a 604-patient study | Huh, W. K. and Cestero, R. M. and Garcia, F. A. and Gold, M. A. and Guido, R. S. and McIntyre-Seltman, K. and Harper, D. M. and Burke, L. and Sum, S. T. and Flewelling, R. F. and Alvarez, R. D. |
| 19 | Diagnosing Cervical Neoplasia in Rural Brazil Using a Mobile Van Equipped with In Vivo Microscopy: A Cluster-Randomized Community Trial | Hunt, B. and Fregnani, Jhtg and Schwarz, R. A. and Pantano, N. and Tesoni, S. and Possati-Resende, J. C. and Antoniazzi, M. and de Oliveira Fonseca, B. and de Macêdo Matsushita, G. and Scapulatempo-Neto, C. and Kerr, L. and Castle, P. E. and Schmeler, K. and Richards-Kortum, R. |
| 20 | A comprehensive study on the multi-class cervical cancer diagnostic prediction on pap smear images using a fusion-based decision from ensemble deep convolutional neural network | Hussain, E. and Mahanta, L. B. and Das, C. R. and Talukdar, R. K. |
| 21 | Image Registration based Cervical Cancer Detection and Segmentation Using ANFIS Classifier | Jaya, B. K. and Kumar, S. S. |
| 22 | Quantitative analysis of abnormalities in gynecologic cytopathology with deep learning | Ke, J. and Shen, Y. and Lu, Y. and Deng, J. and Wright, J. D. and Zhang, Y. and Huang, Q. and Wang, D. and Jing, N. and Liang, X. and Jiang, F. |
| 23 | A study on development of automation diagnosis of liquid based cytology | Kim, S. H. and Oh, H. Y. and Kim, D. W. |
| 24 | Multifeature Quantification of Nuclear Properties from Images of H&amp;E-Stained Biopsy Material for Investigating Changes in Nuclear Structure with Advancing CIN Grade | Konstandinou, C. and Glotsos, D. and Kostopoulos, S. and Kalatzis, I. and Ravazoula, P. and Michail, G. and Lavdas, E. and Cavouras, D. and Sakellaropoulos, G. |
| 25 | Andriod Device-Based Cervical Cancer Screening for Resource-Poor Settings | Kudva, V. and Prasad, K. and Guruvare, S. |
| 26 | Hybrid Transfer Learning for Classification of Uterine Cervix Images for Cervical Cancer Screening. | Kudva, Vidya and Prasad, Keerthana and Guruvare, Shyamala |
| 27 | Detection of Cervical Cancer Cells in Whole Slide Images Using Deformable and Global Context Aware Faster RCNN-FPN | Li, X. and Xu, Z. and Shen, X. and Zhou, Y. and Xiao, B. and Li, T. Q. |
| 28 | Computer-Aided Cervical Cancer Diagnosis Using Time-Lapsed Colposcopic Images | Li, Y. and Chen, J. and Xue, P. and Tang, C. and Chang, J. and Chu, C. and Ma, K. and Li, Q. and Zheng, Y. and Qiao, Y. |
| 29 | Comparison detector for cervical cell/clumps detection in the limited data scenario | Liang, Y. X. and Tang, Z. H. and Yan, M. and Chen, J. L. and Liu, Q. and Xiang, Y. |
| 30 | Dual-path network with synergistic grouping loss and evidence driven risk stratification for whole slide cervical image analysis | Lin, H. and Chen, H. and Wang, X. and Wang, Q. and Wang, L. and Heng, P. A. |
| 31 | Cervical cancer detection in cervical smear images using deep pyramid inference with refinement and spatial-aware booster | Ma, D. and Liu, J. and Li, J. and Zhou, Y. |
| 32 | A fuzzy rank-based ensemble of CNN models for classification of cervical cytology | Manna, A. and Kundu, R. and Kaplun, D. and Sinitca, A. and Sarkar, R. |
| 33 | An effective diagnosis of cervical cancer neoplasia by extracting the diagnostic features using CRF | Mary Pretty , D. and Anandan, V. and Srinivasagan, K. G. |
| 34 | Computer-assisted diagnosis in colposcopy: Results of a preliminary experiment? | Mehlhorn, G. and Mnzenmayer, C. and Benz, M. and Kage, A. and Beckmann, M. W. and Wittenberg, T. |
| 35 | A Cervical Histopathology Dataset for Computer Aided Diagnosis of Precancerous Lesions | Meng, Z. and Zhao, Z. and Li, B. and Su, F. and Guo, L. |
| 36 | A mobile-based image analysis system for cervical cancer detection | Monsur, S. A. and Adeshina, S. A. and Sud, S. and Soboyejo, W. O. |
| 37 | Overlapping Cervical Nuclei Separation using Watershed Transformation and Elliptical Approach in Pap Smear Images. | Muhimmah, Izzati and Kurniawan, Rahadian and Indrayanti |
| 38 | Cervical Cancer Identification Based Texture Analysis Using GLCM-KELM on Colposcopy Data | Novitasari, D. C. R. and Asyhar, A. H. and Thohir, M. and Arifin, A. Z. and Mu'jizah, H. and Foeady, A. Z. |
| 39 | Deep multiple-instance learning for abnormal cell detection in cervical histopathology images | Pal, A. and Xue, Z. and Desai, K. and Aina, F. Banjo A. and Adepiti, C. A. and Long, L. R. and Schiffman, M. and Antani, S. |
| 40 | Domain-specific image analysis for cervical neoplasia detection based on conditional random fields | Park, S. Y. and Sargent, D. and Lieberman, R. and Gustafsson, U. |
| 41 | Comparison of machine and deep learning for the classification of cervical cancer based on cervicography images | Park, Y. R. and Kim, Y. J. and Ju, W. and Nam, K. and Kim, S. and Kim, K. G. |
| 42 | Development of Low-Cost Point-of-Care Technologies for Cervical Cancer Prevention Based on a Single-Board Computer | Parra, S. and Carranza, E. and Coole, J. and Hunt, B. and Smith, C. and Keahey, P. and Maza, M. and Schmeler, K. and Richards-Kortum, R. |
| 43 | Diagnosis of cervical precancerous lesions based on multimodal feature changes | Peng, G. and Dong, H. and Liang, T. and Li, L. and Liu, J. |
| 44 | Real-Time Monitoring and Evaluation of a Visual-Based Cervical Cancer Screening Program Using a Decision Support Job Aid | Peterson, C. W. and Rose, D. and Mink, J. and Levitz, D. |
| 45 | Computerized delineation of nuclei in liquid-based Pap smears stained with immunohistochemical biomarkers | Qin, Y. and Walts, A. E. and Knudsen, B. S. and Gertych, A. |
| 46 | Computer aided decision support system for cervical cancer classification | Rahmadwati, R. and Naghdy, G. and Ros, M. and Todd, C. |
| 47 | Objective screening for cervical cancer in developing nations: lessons from Nigeria | Roblyer, D. and Richards-Kortum, R. and Park, S. Y. and Adewole, I. and Follen, M. |
| 48 | ColpoNet for automated cervical cancer screening using colposcopy images. | Saini, Sumindar Kaur and Bansal, Vasudha and Kaur, Ravinder and Juneja, Mamta |
| 49 | Towards the mobile detection of cervical lesions: A region-based approach for the analysis of microscopic images | Sampaio, A. F. and Rosado, L. and Vasconcelos, M. J. M. |
| 50 | Classification and comparison of malignancy detection of cervical cells based on nucleus and textural features in microscopic images of uterine cervix | Shanthi, P. B. and Modi, S. and Hareesha, K. S. and Kumar, S. |
| 51 | Quantification of confocal fluorescence microscopy for the detection of cervical intraepithelial neoplasia. | Sheikhzadeh, Fahime and Ward, Rabab K and Carraro, Anita and Chen, Zhao Yang and van Niekerk, Dirk and Miller, Dianne and Ehlen, Tom and MacAulay, Calum E and Follen, Michele and Lane, Pierre M and Guillaud, Martial |
| 52 | Classification of colposcopic cervigrams using EMD in R | Shrivastav, K. D. and Mukherjee Das, A. and Singh, H. and Ranjan, P. and Janardhanan, R. |
| 53 | A framework for diagnosing cervical cancer disease based on feedforward MLP neural network and ThinPrep histopathological cell image features. | Sokouti, Babak and Haghipour, Siamak and Tabrizi, Ali |
| 54 | Comparing Deep Learning Models for Multi-cell Classification in Liquid- based Cervical Cytology Image | Sornapudi, S. and Brown, G. T. and Xue, Z. and Long, R. and Allen, L. and Antani, S. |
| 55 | A Unified Model-Based Image Analysis Framework for Automated Detection of Precancerous Lesions in Digitized Uterine Cervix Images | Srinivasan, Y. and Corona, E. and Nutter, B. and Mitra, S. and Bhattacharya, S. |
| 56 | Cervical Cancer Diagnosis based on Random Forest. | Sun Guanglu and Shaobo Li and Yanzhen Cao and Fei Lang |
| 57 | Classification of cervical cancer using Deep Learning Algorithm | Tripathi, A. and Arora, A. and Bhan, A. |
| 58 | Using dynamic features for automatic cervical precancer detection | Viñals, R. and Vassilakos, P. and Rad, M. S. and Undurraga, M. and Petignat, P. and Thiran, J. P. |
| 59 | Artificial intelligence-assisted fast screening cervical high grade squamous intraepithelial lesion and squamous cell carcinoma diagnosis and treatment planning | Wang, C. W. and Liou, Y. A. and Lin, Y. J. and Chang, C. C. and Chu, P. H. and Lee, Y. C. and Wang, C. H. and Chao, T. K. |
| 60 | Automated detection and analysis of fluorescent in situ hybridization spots depicted in digital microscopic images of Pap-smear specimens | Wang, X. W. and Zheng, B. and Li, S. B. and Zhang, R. and Mulvihill, J. J. and Chen, W. R. and Liu, H. |
| 61 | A pap-smear analysis tool (PAT) for detection of cervical cancer from pap-smear images | William, W. and Ware, A. and Basaza-Ejiri, A. H. and Obungoloch, J. |
| 62 | Computer-assisted screening for cervical cancer using digital image processing of pap smear images | Win, K. P. and Kitjaidure, Y. and Hamamoto, K. and Aung, T. M. |
| 63 | Development and validation of an artificial intelligence system for grading colposcopic impressions and guiding biopsies. | Xue, Peng and Tang, Chao and Li, Qing and Li, Yuexiang and Shen, Yu and Zhao, Yuqian and Chen, Jiawei and Wu, Jianrong and Li, Longyu and Wang, Wei and Li, Yucong and Cui, Xiaoli and Zhang, Shaokai and Zhang, Wenhua and Zhang, Xun and Ma, Kai and Zheng, Yefeng and Qian, Tianyi and Ng, Man Tat Alexander and Liu, Zhihua |
| 64 | Prediction Using Hierarchical Data: Applications for Automated Detection of Cervical Cancer | Yamal, J. M. and Guillaud, M. and Atkinson, E. N. and Follen, M. and MacAulay, C. and Cantor, S. B. and Cox, D. D. |
| 65 | HLDnet: Novel deep learning based Artificial Intelligence tool fuses acetic acid and Lugol's iodine cervicograms for accurate pre-cancer screening | Yan, L. and Song, H. and Guo, Y. and Ren, P. and Zhou, W. and Li, S. and Yang, J. and Shen, X. |
| 66 | Contrast-Enhancing Snapshot Narrow-Band Imaging Method for Real-Time Computer-Aided Cervical Cancer Screening. | Yi, Dingrong and Kong, Linghua and Zhao, Yanli |
| 67 | The application of deep learning based diagnostic system to cervical squamous intraepithelial lesions recognition in colposcopy images | Yuan, C. and Yao, Y. and Cheng, B. and Cheng, Y. and Li, Y. and Li, Y. and Liu, X. and Cheng, X. and Xie, X. and Wu, J. and Wang, X. and Lu, W. |
| 68 | Automation-assisted cervical cancer screening in manual liquid-based cytology with hematoxylin and eosin staining | Zhang, L. and Kong, H. and Ting Chin, C. and Liu, S. and Fan, X. and Wang, T. and Chen, S. |
| 69 | Automatic screening of cervical cells using block image processing | Zhao, M. and Wu, A. and Song, J. and Sun, X. and Dong, N. |
| 70 | Hybrid AI-assistive diagnostic model permits rapid TBS classification of cervical liquid-based thin-layer cell smears | Zhu, X. and Li, X. and Ong, K. and Zhang, W. and Li, W. and Li, L. and Young, D. and Su, Y. and Shang, B. and Peng, L. and Xiong, W. and Liu, Y. and Liao, W. and Xu, J. and Wang, F. and Liao, Q. and Li, S. and Liao, M. and Li, Y. and Rao, L. and Lin, J. and Shi, J. and You, Z. and Zhong, W. and Liang, X. and Han, H. and Zhang, Y. and Tang, N. and Hu, A. and Gao, H. and Cheng, Z. and Liang, L. and Yu, W. and Ding, Y. |
| 71 | Automation of detection of cervical cancer using convolutional neural networks | Kudva, V. and Prasad, K. and Guruvare, S. |
| 72 | Towards rapid cervical cancer diagnosis: automated detection and classification of pathologic cells in phase-contrast images | Schilling, T., Miroslaw, L., Glab, G. & Smereka, M. |
| 73 | Intelligent machine learning based computer aided diagnosis model for cervical cancer detection and classification | Suguna, C., Balamurugan, S. P. |

Supplementary Table 3: Summary of novel automated cervical cancer screening devices and their development stages

| **Medical Device** | **Screening Domain** | **Author** | **Study objective** | **Stage of development of medical device (lifecycle)** |
| --- | --- | --- | --- | --- |
| Pocket Colposcope | Colposcopy | Asiedu et al. (2018) | To propose methods automatic classification and methods for combining features of different contrasts for improved performance.  An **SVM classifier** was used. | **Verification & validation phase** (pre-market clinical trials). Device commercialized but not algorithm |
| MobileODT EVA Colpo | Colposcopy | Xue et al. (2020a) | To report evaluation of AVE performance on EVA Colpo-acquired images.  A **Faster R-CNN** model was used. | **Verification & validation phase** (pre-market clinical trials). Device commercialized but not algorithm |
| MobileODT EVA Colpo | Colposcopy | Peterson et al. (2016) | To test the implementation of the novel job aid feature on the mobile colposcope application during an EVA deployment in Kenya.  The model name was not disclosed. | **Verification & validation phase** (pre-market clinical trials). Device commercialized but not algorithm |
| MobileODT EVA Colpo | Colposcopy | Chandran et al. (2021) | To present a computerised system for cervical cancer prediction from colposcopy images. A **CYENET** model was used. | **Verification & validation phase** (pre-market clinical trials). Device commercialized but not algorithm |
| Dr Cervicam C20 | Colposcopy | Park et al. (2021) | To conduct comparative binary classification experiments using machine learning and deep learning techniques in the same environment and assess best performing classifiers.  The highest performing ML model was **SVM classifier**. **ResNet-50** was the deep learning model used. | **Verification & validation phase** (pre-market clinical trials). Device commercialized but not algorithm |
| Smartphone camera | Colposcopy | Vinals et al. (2021) | To proposes a smartphone-based solution that automatically detects cervical precancer from the dynamic features extracted from videos taken during VIA. An **ANN model** was used. | **Feasibility phase**: Device is commercially available, however, not for the purposes proposed**. Algorithm is in feasibility stage. |
| Smartphone camera | Colposcopy | Kudva et al. (2018a) | To propose an algorithm for analysis of cervix images acquired using an Android™ device, which can be used for the development of decision support system. An **SVM classifier** was used. | **Feasibility phase**: Device is commercially available, however, not for the purposes proposed**. Algorithm is in feasibility stage. |
| Smartphone camera | Colposcopy | Kudva et al. (2018b) | To propose an analytic procedure that evaluates cervix images acquired using an Android device and utilizes convolutional neural networks which achieve 100% accuracy in classifying cervix images. A shallow **CNN** model was used. | **Feasibility phase**: Device is commercially available, however, not for the purposes proposed**. Algorithm is in feasibility stage. |
| Smartphone camera | Colposcopy | Monsur et al. (2017) | To propose a method for integration and automation of several complex image processing and computer vision techniques into a mobile device using low quality images. **Mean gray values** was used. | **Feasibility phase**: Device is commercially available, however, not for the purposes proposed**. Algorithm is in feasibility stage. |
| Smartphone-based endoscope system | Colposcopy | Bae et al. (2020) | To demonstrate a new quantitative cervical cancer screening technique and implement a machine learning algorithm for smart phone-based endoscopic VIA. A **KNN** model was used. | **Feasibility phase**: Device and algorithm not distributed in the market. |
| Grandium Ocus | Cytology | Holmstrom et al. (2021) | To determine whether artificial intelligence–supported digital microscopy diagnostics can be implemented in a resource-limited setting and used for analysis of Papanicolaou ‘pap’ smear tests. A **CNN** model was used. | **Product launch**: Regulator approved or Commercially available. |
| Cytosavant system | Cytology | Yamal et al. (2015) | To perform a comparative study of three main approaches for problems with hierarchical data structure: a) extract patient-level features from the cell-level data; b) use a statistical model that accounts for the hierarchical data structure; and c) classify at the cellular level, and use an ad hoc approach to classify at the patient level. **Decision trees** model was used. | **Feasibility phase**: Device and algorithm not on the market |
| SmartScope | Cytology | Sampaio et al. (2021) | To investigate the possibility of transferring domain knowledge by training CNN models with a public dataset closer to application domain then fine-tuning models with dataset generated by novel device under investigation. An **F-RCNN** model was used. | **Verification & validation phase** (pre-market clinical trials) |
| FISH imaging microscope | Cytology | Wang et al. (2009) | To develop and test a computerized scheme aimed to more reliably and robustly detect analysable interphase cells and to analyse related FISH spots. A **blue colour space histogram** was used. | **Feasibility phase**: Device and algorithm not on the market |
| J5 samsung smartphone affixed to microscope | Histopathology | Pal et al. (2021) | To assess the feasibility of developing a low-cost diagnostic system with the H&E stained cervical tissue image analysis algorithm. A **Deep-MIL** model was used. | **Feasibility phase**: Device and algorithm not on the market |
| mHRME | Micro-endoscopy | Hunt et al. (2018) | To compare rates of diagnostic follow-up completion between two study arms of receiving medical care at mobile clinic vs. at central hospital; and to evaluate the diagnostic performance of in vivo microscopy compared with colposcopy. Model used was not disclosed. | **Feasibility phase**: Device and algorithm not on the market |
| PiHRME | Micro-endoscopy | Parra et al. (2020) | To demonstrate a low-cost imaging system for real-time detection of cervical precancer that uses a single-board computer. A **MobileNetV2** model was used. | **Feasibility phase**: Device and algorithm not on the market |
